# COVID-19 increases age- and sex-controlled 21-day fatality rates for patients with melanoma, hematologic malignancies, uterine cancer, or kidney cancer

**DOI:** 10.1101/2021.02.06.21251099

**Authors:** Haiquan Li, Edwin Baldwin, Xiang Zhang, Colleen Kenost, Wenting Luo, Elizabeth A. Calhoun, Lingling An, Charles L. Bennett, Yves A. Lussier

## Abstract

**Introduction:** Prior research has reported an increased risk of fatality for cancer patients, but most studies investigated the risk by comparing cancer patients to non-cancer patients among COVID-19 infections. Only a few studies have compared the impact of a COVID-19 infection to non-infection with matched cancer patients and types.

**Methods & Materials:** We conducted survival analyses of 4,606 cancer patients with COVID-19 test results from March 16 to October 11, 2020 in UK Biobank and estimated the overall hazard ratio of fatality with and without COVID-19 infection. We also examined the hazard ratios of thirteen specific cancer types with at least 100 patients.

**Results:** COVID-19 resulted in an overall hazard ratio of 7.76 (95% CI: [5.78, 10.40], p<10^−10^) by studying the survival rate of 4,606 cancer patients for 21-days after the tests. The hazard ratio was shown to vary among cancer type, with over a 10-fold increase in fatality rate (false discovery rate≤0.02) for melanoma, hematologic malignancies, uterine cancer, and kidney cancer using a stratified analysis on each of the cancer types. Although COVID-19 imposed a higher risk for localized cancers compared to distant metastasis ones, those of distant metastasis yielded higher fatality rates due to their multiplicative effects.

**Conclusion:** The results highlight the importance of timely care for localized and hematological cancer patients and the necessity to vaccinate uninfected patients as soon as possible, particularly for the cancer types influenced most by COVID-19.

## Introduction

For persons with localized cancers or hematologic malignancies, timely cancer treatment is critical^1^. However, with COVID-19 markedly impacting cancer care, treatment delays are inevitable^2,3^. Therefore, attention to the timeliness of therapy for patients with cancer is encouraged^4^. Still, the extent of risk that delays in cancer therapies add for persons with COVID-19 is not known^5,6^ and is likely to vary depending on cancer type ^7^, stage, grade, and treatment^6,8^. Although prior research has been conducted, most studied the hazard ratio (HR) or odds ratio by comparing cancer patients to non-cancer patients among COVID-19 patients, which did not reflect the impact of COVID-19 on specific cancer types^9-11^. Passamonti et al. reported 536 hematological cancer patients with COVID-19 infection at 66 hospitals in Italy^12^. Those patients with severe or critical COVID-19 infections had a HR of 4.08 for mortality compared to those with mild severity in Italy^12^. We report a study comparing fatality rates among persons with a wide range of cancer diagnoses with and without COVID-19 while controlling for age, sex, and type of cancer.

## Methods

We conducted a retrospective survival study using UK Biobank (UKB) under the UKB COVID-19 policy. UKB is a biobank with longitudinal COVID-19 test results, death registries, cancer registries, and inpatient records for approximately 500,000 patients. COVID-19 tests started on March 16, 2020 for symptomatic patients, during which testing capacity was limited, and results were provided by Public Health England^13^. There were roughly 67,000 living cancer subjects at the beginning of the test. Inclusion criteria for the study were (i)subjects of British Ancestry with a past history of hospitalization in UKB (updated to the end of September 2020) and (ii) subjects conducted a COVID-19 test no later than October 11, 2020, for which we could obtain a 21-day follow up in the death registry from NHS Digital and NHS Central Register, UK. That left 6,528 cancer patients. We excluded 893 cancer patients whose cancer diagnoses were ten or more years ago, without another primary cancer or recurrence in the record since they are unlikely in remission and more closely resemble non-cancer patients. We further excluded five cancer patients with inconsistent self-report of sex and 1,024 “non-melanoma skin cancer patients” reported with truncated ICD-10 codes in the UKB, which conflates non-lethal basal cell carcinomas with lethal forms of non-melanoma skin cancers. The final cancer cohort comprised 4,606 patients, where 288 (6.3%) were positive for COVID-19 (**Table 1**). We also built a randomized non-cancer cohort of 4,606 patients for comparative studies, which matched the COVID-19 status, sex, age (per 5-year bin), and specific laboratory testing facility of the corresponding cancer patients. We exclude seven non-cancer patients with COVID-19 tests conducted after death during the sampling.

**Table 1:** Demographic and Clinical characteristics of 4,606 COVID-19 tested cancer subjects in the UK Biobank.

|  | COVID-19 positive-associated |  |  | COVID-19 negative-associated |  |  |
| --- | --- | --- | --- | --- | --- | --- |
|  | Fatalities | (%) | Survivors | Fatalities | (%) | Survivors |
| **Race** |  |  |  |  |  |  |
| British Ancestry | 64 | (22.2) | 224 | 153 | (3.5) | 4318 |
| **Age** |  |  |  |  |  |  |
| 50-59 | 2 | (7.1) | 26 | 8 | (2.9) | 271 |
| 60-69 | 6 | (9.7) | 56 | 26 | (3.0) | 850 |
| 70-79 | 42 | (26.4) | 117 | 94 | (3.5) | 2562 |
| 80-84 | 14 | (35.9) | 25 | 25 | (4.9) | 482 |
| **Sex** |  |  |  |  |  |  |
| Male | 40 | (23.8) | 128 | 90 | (3.8) | 2285 |
| Female | 24 | (20.0) | 96 | 63 | (3.2) | 1880 |
| **History of cancer** |  |  |  |  |  |  |
| yrs > 10 | 18 | (27.7) | 47 | 35 | (3.4) | 1008 |
| 5 <yrs ≤ 10 | 15 | (18.5) | 66 | 24 | (2.3) | 1005 |
| 1 <yrs ≤ 5 | 3 | (13.0) | 20 | 8 | (4.3) | 178 |
| yrs ≤ 1 | 28 | (23.5) | 91 | 86 | (4.2) | 1974 |

We analyzed the case fatality rate (CFR) of cancer patients and their cancer types using a multivariate Cox proportional hazard model. COVID-19 status was obtained from the patient’s first test result (negative controls remained so throughout). Control covariates included ethnicity, age, sex, and cancer status. Age was treated as a continuous variable and cancer status was categorized as localized versus metastatic based on UKB ICD-10 codes from inpatient records. R’s *survival* package was used to diagnose the model and plot Kaplan Meier curves.

## Results

Quartiles of time from COVID-19 diagnosis to death for cancer patients (64 total) are 5, 10, and 14 days, respectively. The CFR within 21 days of diagnosis for COVID-19 positive cancer patients was six-fold higher than that of COVID-19 negative ones, 22.2% (64 out of 288) versus 3.5% (153 out of 4,318) (**Table 1**). Distant metastasis patients demonstrated a higher fatality rate, where CFR within 21 days of a positive COVID-19 diagnosis was nearly double that of localized cancer patients, 37.7% versus 18.7% (**Figure 1a & 1c & 1d**). No matter the spread of cancer, the CFR of COVID-19 within 21 days (22.2%) was higher than that of non-cancer patients when positive (15.6%), from the cohort using matched covariates such as test result, sex, age, and testing venue (a proxy of the hospitalization system). A multivariate Cox model was built to study individual risk factors and showed a HR of 7.76 (95% CI: [5.78, 10.40], p<10^−10^) for COVID-19 positive cancer patients compared to COVID-19 negative cancer patients after controlling for ethnicity, sex, age, and metastatic versus localized cancer confounders (Figure 1). The model suggested increased fatality rates by COVID-19 infections particularly for patients with melanoma, lymphoma, leukemia, uterine, or kidney cancer. The HRs of COVID-19 were 10-fold higher for these five cancer diagnoses using stratified analyses with matched cancer types **(Figure 1)**. As expected, distant metastasis was a risk factor for fatality, with a HR of 3.92 (95% CI: [2.99, 5.13]; p<10^−10^) compared to localized cancers. Age remained an important factor, with a HR of 1.04 per year (95% CI:1.02-1.07; p=3.1×10^−4^), but sex was not significant. No factor was found to significantly deviate from the model assumption (p>0.25). Logistic regression led to similar but slightly inflated results.

**Figure 1.**
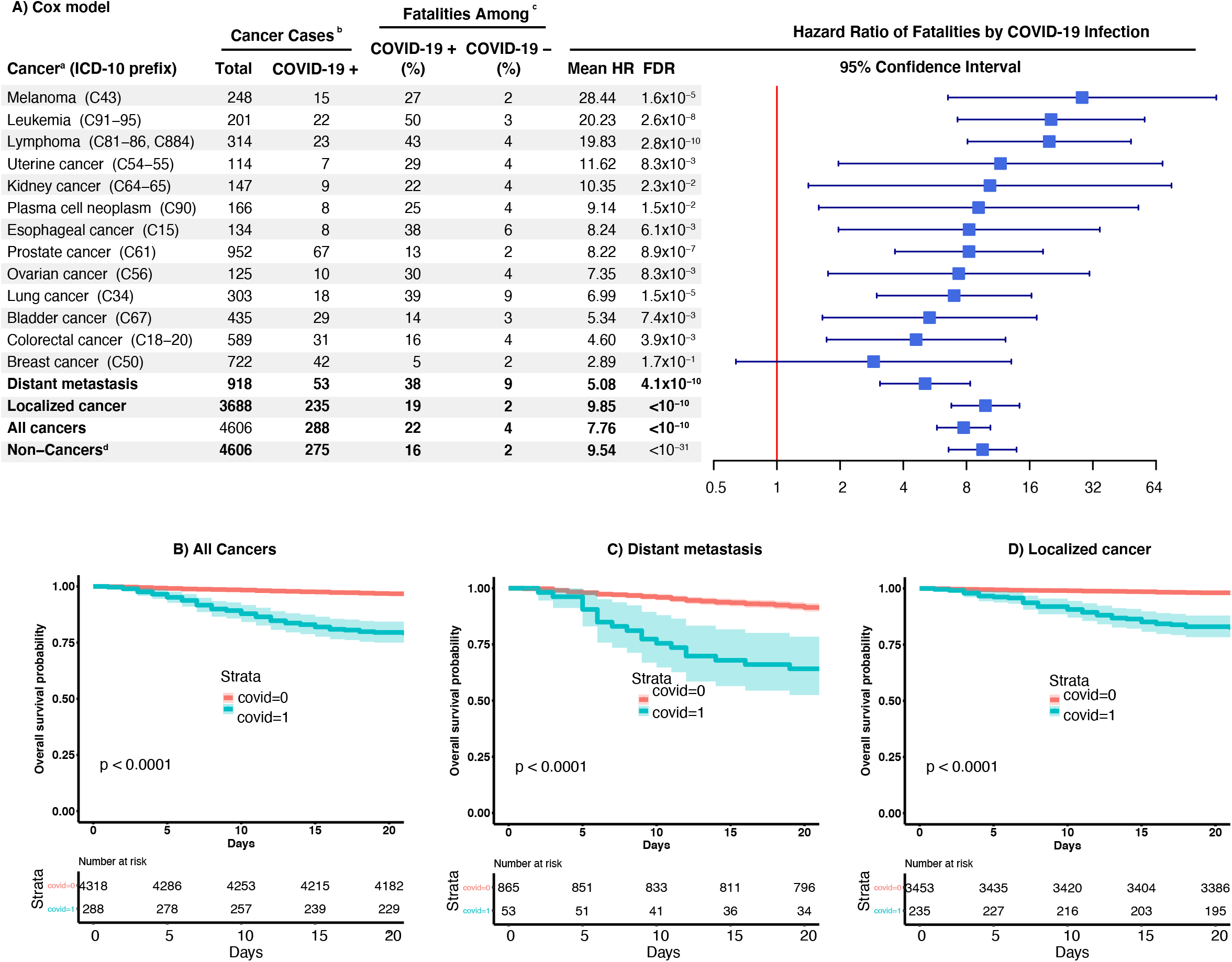
Hazard ratios of COVID-19-associated death among cancer patients and types of cancers. a) Results were from stratified analyses using the patients stratifying the conditions listed in the column named ‘cancer’. All analyses used Cox model with covariates of sex, age, COVID-19 infection status, cancer status (localized or distant metastasis) excepted stratified analyses of localized cancer and distant metastasis. Cancer subtypes were analyzed when they were composed of 100 or more total cases. b) Four cancer patients with inconsistent self-reporting and gene sex were excluded from the study. c) Fatality event was assessed for the 21 days following the first COVID-19 positive testing or the first COVID-19 negative testing (negative controls remained so throughout) and was available for all cancer subjects under study. d) Sixty non-cancer patients did not match the testing facility with a cancer patient due to lacking subjects of matching all factors (e.g., age).

## Discussion

Our study demonstrated that COVID-19 adds 10-fold more risk to 21-day fatality rates for patients with melanoma, lymphoma, leukemia, uterine, and kidney cancer with a positive COVID-19 infection versus no COVID-19 infection. Our results for lymphoma and leukemia patients confirm reports from Italy^12^, while the findings of increased 21-day mortality rates in COVID-19 infections among melanoma, uterine, and kidney cancer patients have not been reported previously. Fortunately, our study suggests that COVID-19 does not impose larger risk to distant metastasized cancers as compared to localized cancers (e.g., lymphoma, leukemia) in general. However, the overall fatality rate in distant metastases was still about twice that of localized cancers due to the multiplicative effect of hazard ratios in our model. It should be noted that fatality rates were dependent on cancer type and COVID-19 did impose larger risk to distant metastasis of some types of cancer, such as melanoma (HR 49.37, 95% CI: [3.70, 658.85]), prostate cancer (HR 22.11, 95% CI: [6.15, 79.54]), and ovarian cancer (HR 13.04, 95% CI: [2.61,65.14]), based on a stratified analysis using only metastasis patients (Supplemental Figure 1). In all cases, higher rates of fatality among COVID-19 patients of older age were consistent with the literature. The strengths of this study include the large UKB cancer cohort size for COVID-19 patients and its reliable death registry. Limitations include the unavailability of cancer stage and grade plus a relatively small sample size for some specific cancer types. Further, the study was unable to include other preexisting conditions that may have been associated with the fatality, and the conclusions may be limited to symptomatic patients and hospitalized patients due to the inclusion criteria. Our findings reinforce the clinical importance of timely treatment of COVID-19 among older cancer patients with hematological malignancies, melanoma, uterine, or kidney cancer. The findings also support specific guidelines emphasizing the importance of timely care for COVID-19 infected persons and strongly support a change in COVID-19 vaccine strategy with hematologic malignancies in particular.

## Data Availability

The results will be shared upon request. The original data was from UK Biobank so the access should follow UK Biobank data access application process.

## Acknowledgments

We have no conflicts of interest to disclose.

