## Supplemental Figure 1 for "COVID-19 increases age- and sex-controlled 21-day fatality rates for patients with melanoma, hematologic malignancies, uterine cancer, or kidney cancer"

| Cancer* (ICD-10 prefix) | Cancer Cases: Total | Cancer Cases: COVID-19+ | Fatalities Among COVID-19+ (%) | Fatalities Among COVID-19– (%) | Mean HR | HR FDR |
|---|---|---|---|---|---|---|
| **Localized cancer** | 3688 | 235 | 19 | 2 | 9.85 | $5.9 \times 10^{-32}$ |
| Lymphoma (C81–86,C884) | 292 | 20 | 45 | 4.04 | 21.28 | $1.0 \times 10^{-9}$ |
| Melanoma (C43) | 206 | 13 | 15 | 1.04 | 20.31 | $9.2 \times 10^{-3}$ |
| Leukemia (C91–95) | 192 | 22 | 50 | 3.53 | 20.23 | $3.7 \times 10^{-8}$ |
| Plasma cell neoplasm (C90) | 151 | 7 | 29 | 3.47 | 14.25 | $1.3 \times 10^{-2}$ |
| Colorectal cancer (C18–20) | 431 | 25 | 20 | 1.72 | 12.04 | $6.2 \times 10^{-5}$ |
| Breast cancer (C50) | 594 | 36 | 6 | 0.54 | 10.21 | $1.4 \times 10^{-2}$ |
| Kidney cancer (C64–65) | 110 | 9 | 22 | 1.98 | 8.84 | $3.7 \times 10^{-2}$ |
| Lung cancer (C34) | 198 | 12 | 33 | 5.91 | 6.57 | $3.5 \times 10^{-3}$ |
| Prostate cancer (C61) | 816 | 59 | 7 | 1.59 | 4.20 | $1.5 \times 10^{-2}$ |
| Bladder cancer (C67) | 399 | 26 | 8 | 2.14 | 4.07 | $7.8 \times 10^{-2}$ |
| **Distant metastasis** | 918 | 53 | 38 | 9 | 5.08 | $9.6 \times 10^{-10}$ |
| Melanoma (C43) | 42 | 2 | 100 | 8 | 49.37 | $5.7 \times 10^{-3}$ |
| Prostate cancer (C61) | 136 | 8 | 62 | 4 | 22.11 | $6.8 \times 10^{-6}$ |
| Ovarian cancer (C56) | 91 | 7 | 43 | 4 | 13.04 | $3.5 \times 10^{-3}$ |
| Lung cancer (C34) | 105 | 6 | 50 | 15 | 5.95 | $1.3 \times 10^{-2}$ |

**Hazard Ratio of COVID-19 Fatalities**

**95% Confidence Interval**

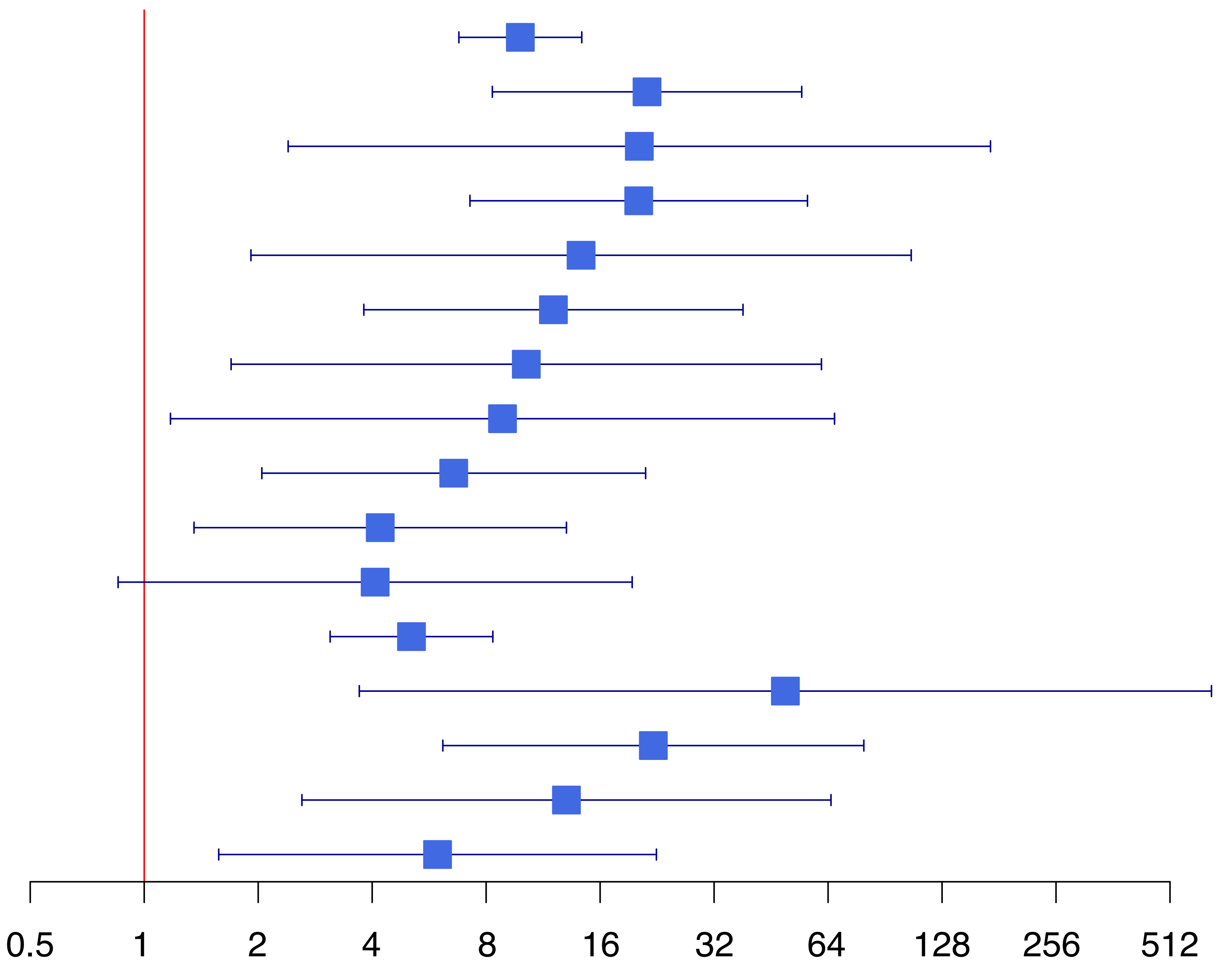


Notes:
1) All stratiffied analyses use COVID-19 plus covariates sex and age.
2) Localized cancer type stratified studies included those with at least 100 localized cases.
3) Distant metastsis cancer type stratified studies included those with at lesat 40 metastasized cases, except colorectal, breast, and panncreatic cancers, which had no COVID-19 associated deaths.
